# Persistence of immunity against omicron BA.1 and BA.2 following homologous and heterologous COVID-19 booster vaccines in healthy adults after a two-doses AZD1222 vaccination

**DOI:** 10.1101/2022.06.05.22276016

**Authors:** Suvichada Assawakosri, Sitthichai Kanokudom, Jira Chansaenroj, Nungruthai Suntronwong, Chompoonut Auphimai, Pornjarim Nilyanimit, Preeyaporn Vichaiwattana, Thanunrat Thongmee, Thaneeya Duangchinda, Warangkana Chantima, Pattarakul Pakchotanon, Donchida Srimuan, Thaksaporn Thatsanatorn, Sirapa Klinfueng, Natthinee Sudhinaraset, Nasamon Wanlapakorn, Juthathip Mongkolsapaya, Sittisak Honsawek, Yong Poovorawan

## Abstract

**Objectives:** The SARS-CoV-2 Omicron variant presents numerous mutations potentially able to evade neutralizing antibodies (NAbs) elicited by COVID-19 vaccines. Therefore, this study aimed to provide evidence on a heterologous booster strategy to overcome the waning immunity against Omicron variants.

**Methods:** Participants who completed Oxford/AstraZeneca (hereafter AZD1222) for 5-7 months were enrolled. The reactogenicity and persistence of immunogenicity in both humoral and cellular response after a homologous or heterologous booster with the AZD1222 and mRNA vaccines (BNT162B2, full or half-dose mRNA-1273) administered six months after primary vaccination were determined.

**Results:** Total 229 individuals enrolled, a waning of immunity was observed 5-7 months after the AZD1222-primed. Total RBD immunoglobulin (Ig) levels, anti-RBD IgG and focus reduction neutralization test against Omicron BA.1 and BA.2 and T cell response peaked 14-28 days after booster vaccination. Both the full and half dose of mRNA-1273 induced the highest response, followed by BNT162b2 and AZD1222. At 90 days, the persistence of immunogenicity was observed among all mRNA-boosted individuals. Adverse events were acceptable and well tolerated for all vaccines.

**Conclusions:** A heterologous mRNA booster provided a significantly superior boost of binding and NAbs levels against the Omicron variant compared to a homologous booster in individuals with AZD1222-primed vaccinations.

## Introduction

Since the first emergence of the severe acute respiratory syndrome coronavirus 2 (SARS-CoV-2) Omicron (BA.1/B.1.1.529) variant in November 2021, it has rapidly spread and became the dominant variant circulating worldwide (WHO, 2022a, WHO HQ, 2022). The Omicron variant harbors mutations within the Spike (S) protein, particularly 15 amino acid substitutions in the receptor-binding domain (RBD) (Viana et al., 2022). Mutations within the RBD of the omicron variant mediate antibody evasion and greatly increase transmissibility through enhanced affinity for the angiotensin-converting enzyme 2 receptor (ACE2) (Mannar et al., 2022, McCallum et al., 2022, Tian et al., 2021). Recently, the Omicron variant was further classified into several descendent sublineages, including BA.1, BA.1.1, BA.2, BA.2.2, and BA.3. (Viana et al., 2022). As of March 2022, epidemiological data have suggested that BA.2 sublineages have been the most common sublineage of Omicron worldwide, including the South-East Asia region (WHO, 2022b). Furthermore, like Thailand, epidemiological surveillance revealed that the proportion of the BA.2 variant has increased and represented >90% of all positive cases reported since March 2022 (Puenpa et al., 2022). BA.1 and BA.2 share multiple mutations, but BA.2 also presents unique viral characteristics of BA.1, such as higher reproduction rate, fusogenicity, and pathogenicity (Viana et al., 2022, Yamasoba et al., 2022). Scientific knowledge on the difference in ability between BA.1 and BA.2 to evade third dose vaccine-induced immunity is still limited.

Several coronavirus diseases 2019 (COVID-19) vaccines have been developed to combat the COVID-19 infection. The AZD1222 vaccine has been the highly used vaccine accounting for 2.8 billion doses administered worldwide and 48.3 million doses in Thailand alone (AstraZeneca PLC, 2022, MOPH, 2022). Although the vaccine effectiveness (VE) after two-dose AZD1222 was estimated at 64%–74.0% for preventing SARS-CoV-2 and other lineage infection (Clemens et al., 2021, Falsey et al., 2021), waning of vaccine-induced immunity of both anti-IgG and the NAbs from AZD1222-primed vaccinees has been documented. For example, the dramatically decreased levels of NAbs after 5-6 months of two doses of AZD1222 vaccination showed an inadequate response to control the spread of SARS-CoV-2, especially the Omicron variant (Dejnirattisai et al., 2022, Planas et al., 2022, Shrotri et al., 2021). Like a previous study, NAbs decreased substantially after six months of vaccination with BNT162b2 (Dejnirattisai et al., 2022). Furthermore, the increase in infection was related to the waning immunity as a function of time after BNT162b2-primig (Goldberg et al., 2021, Levin et al., 2021). Thus, the decline in vaccine-induced immunity markedly increased a few months after vaccination and may be the potential cause of breakthrough infection.

There is a growing interest in using booster dose as a new strategy to combat waning immunity after primary vaccination and the high transmissibility and immune evasion of the omicron sublineages. To date, increasing evidence has supported that the booster dose of the mRNA vaccine following BNT162b2-primed significantly increased protection against the Omicron variant (Garcia-Beltran et al., 2022). Furthermore, a heterologous booster after six months of two-dose CoronaVac demonstrated that a heterologous booster produced stronger immunity than a homologous booster (Assawakosri et al., 2022). In individuals primed with AZD1222, a heterologous boost with BNT162b2 exhibited more effectiveness in inducing NAbs against BA.1 than the homologous booster with AZD1222 (Dejnirattisai et al., 2022), but there were no data against BA.2. These results implied that the heterologous booster strategy could provide stronger immunity against Omicron infection. However, very limited knowledge is available on the duration of immune protection after the third homologous or heterologous dose and when we should consider an additional booster beyond the third dose.

Consequently, our objective was to study the persistence of immunogenicity beyond one month following the third dose. In the same cohort, we also investigated NAb against the new emerging omicron variant, particularly the BA.2 sublineage, the cellular response and the reactogenicity after the homologous and heterologous booster dose administered at 6-month intervals in healthy adults who were previously vaccinated with AZD1222.

## MATERIALS AND METHODS

### Study design

This prospective cohort study was conducted at the Clinical Trial Unit, the Center of Excellence in Clinical Virology of Chulalongkorn University, Bangkok, Thailand, and was performed in accordance with the principles of the Declaration of Helsinki and the Good Clinical Practice Guidelines (ICH-GCP). The participants were informed, and written consent was obtained before enrollment.

### Study participants

Between November 2021 and January 2022, a total of 229 subjects participated in this prospective cohort study. The main inclusion criteria were: Thai adults over 18 years of age with no previous or current diagnosis of COVID-19 infection and completed a two-dose vaccination of the AZD1222 at an interval of 5-7 months during the enrollment period. The intervals between two-dose AZD1222-primed were 8-10 weeks. In addition, participants with an autoimmune or cancer disease, taking immunosuppressive drugs, or participants that were pregnant or breastfeeding were excluded.

### Study vaccine

All participants were divided into four groups by conveniently sampling 50-60 participants per group based on vaccine availability and vaccinated with one of the following preparations; including 1) viral vector: AZD1222 (AstraZeneca, Oxford, UK) (Folegatti et al., 2020), 2) mRNA: BNT162b2 (Pfizer-BioNTech Inc., New York City, NY), 3) mRNA: 100 μg mRNA-1273 (full-dose group) (Moderna Inc., Cambridge, MA), and 4) 50 μg mRNA-1273 (half-dose group) (Jackson et al., 2020). The study flow is illustrated in Figure 1.

**Figure 1.**
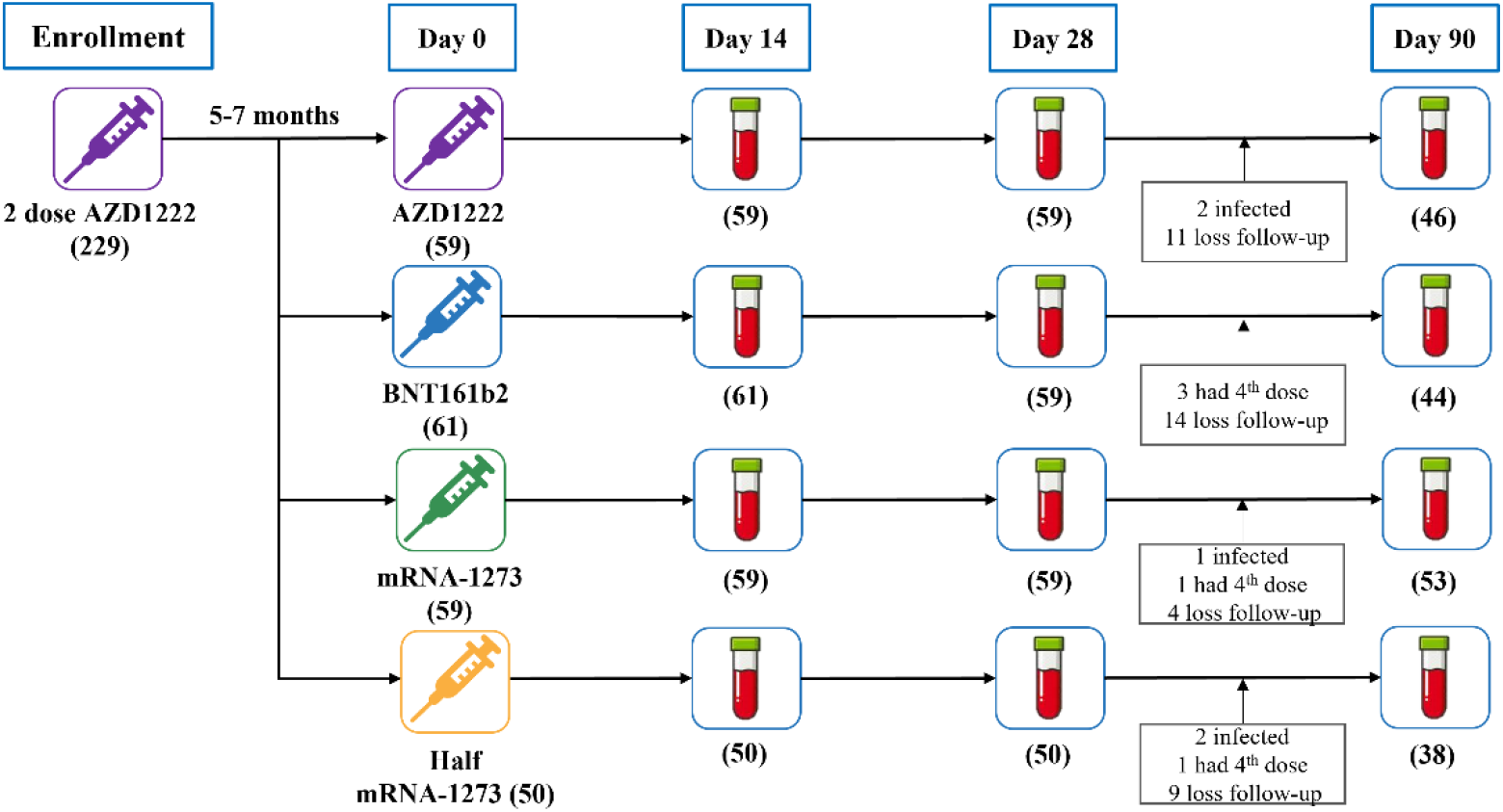
Diagram of the number of included participants and study groups. A total of 229 individuals who were previously vaccinated with AZD1222 were enrolled to analyze binding antibodies and T cell responses in the cohort study. Blood samples were collected at 0, 14, 28, and 90 days after booster vaccination. They were assigned to receive a booster vaccine including AZD1222 (n=59) or BNT162b2 (n=61) or mRNA-1273 (n=59) or half-dose mRNA-1273 (n=50).

### Reactogenicity Assessment

All participants were observed under the supervision of doctors after vaccination to prevent anaphylaxis reactions. Participants received an online or paper-based self-monitoring record survey seven days after vaccination. They observed the solicited local and systemic AEs, including injection site pain, induration, redness, fever, headache, myalgia, arthralgia, nausea, and vomiting.

### Sample collection, total RBD Ig, and anti-RBD IgG/nucleocapsid assay

Blood samples from vaccinees were collected prior to vaccination and at every follow-up visit on days 14, 28, and 90 after the booster dose. The samples of all participants were tested for quantitative antibody levels of total RBD Ig using Elecsys SARS-CoV-2 S (Roche Diagnostics, Basel, Switzerland), anti-RBD IgG, and anti-nucleocapsid IgG (anti-N IgG) using the SARS-CoV-2 IgG assay (Abbott, Sligo, Ireland) (Kanokudom et al., 2022).

### Surrogate virus neutralization assay for delta and omicron variants

A cPass™ SARS-CoV-2 surrogate virus neutralization test (GenScript Biotech, New Jersey) was used to measure NAb titters against SARS-CoV-2 variants. Recombinant RBD from delta and omicron strains and 96-well plates coated with recombinant human ACE2 was used (Kanokudom et al., 2022). The percentage of inhibition of a sample was calculated, samples with percent inhibition ≥ 30% were considered a “seropositive” for SARS-CoV-2 neutralizing antibodies.

### Focus reduction neutralization test (FRNT50)

The neutralization antibody titers against the variants BA.1 and BA.2 of SARS-CoV-2 Omicron were measured. The Focus reduction neutralization assay was performed (Assawakosri et al., 2022). The nucleotide sequences of the BA.1 and BA.2 variants are deposited in GenBank under accession numbers: EPI_ISL_8547017 for BA.1 and EPI_ISL_11698090 for BA.2 variant. The 50% focus reduction percentage was calculated, and the inhibitory concentration (IC50) was determined using PROBIT software. The detection limit of the assay is 1:20, and the NAbs values below the limit were substituted with a titer of 10.

### Interferon-gamma release assay

The interferon-gamma (IFN-γ) release assay was performed to evaluate cell-mediated immunity using QuantiFERON (QFN) SARS-CoV-2 RUO (Qiagen, Hilden, Germany) (Kanokudom et al., 2022). Briefly, heparinized whole blood was added to a spike-peptides-coated tube (Ag1) to stimulate CD4+ and (Ag2) to stimulate CD4+, and CD8+ T cells then incubated for 21 hours. Plasma samples were then collected to measure IFN-γ levels by ELISA (Munro et al.). The IFN-γ levels were expressed in IU IFN-γ/mL. Levels above 0.15 IU/mL were defined as seropositive.

### Statistical analysis

G*power software version 3.1.9.6 was used to calculate the sample size. All statistical analyses were conducted using the Statistical Package for the Social Sciences (SPSS) v.22 (SPSS Inc., Chicago, IL, USA). Figures were generated using GraphPad Prism v9.0 (GraphPad Software, San Diego, CA, USA) and R v4.1.2. Software (R Foundation for Statistical Computing, Vienna, Austria). Binding antibody and NAbs were logarithmically transformed, and comparison between groups was performed using analysis of variance (Folegatti et al.) with Bonferroni adjustment or Kruskal-Wallis H test for nonparametric data. The qualitative data comparison was performed using Pearson’s χ^2^ or Fisher’s exact test. The Wilcoxon signed rank test was used for pairwise analysis. A *p*-value <0.05 was considered statistically significant.

## RESULTS

### Demographic data

From November 2021 to January 2022, 229 adults were enrolled in the study. All participants were healthy Thai adults with a mean age of 50.05 (±9.71) years, 55.72 (±13.68) years, 50.66 (±12.95) years, and 57.02 (±12.28) years in group AZD1222, BNT162b2, full-dose mRNA-1273 and half-dose mRNA-1273, respectively. Most participants were female, 127 (55.5%) participants, and the average interval between the second dose and the booster dose was 166.3 (128-229) days. Common comorbidities included dyslipidemia, hypertension, diabetes mellitus, and allergy. Generally, the groups did not have significant differences in baseline characteristics, including sex and common underlying diseases and the intervals between follow-up times.

However, the age of BNT162b2 and half-dose mRNA-1273 was significantly higher than that of AZD1222 and mRNA-1273 arms. Participant characteristics are shown in Table 1. During the study, 38 participants were lost to follow-up, five participants were excluded because they tested positive for SARS-CoV-2 infection, and another five participants were excluded because they had received a fourth vaccine dose.

**Table 1:** Baseline characteristics of the participants enrolled in this study.

|  | **Total** | **AZD1222** | **BNT162b2** | **mRNA-1273** | **Half mRNA-1273** |
| --- | --- | --- | --- | --- | --- |
| **Total number (n)** | 229 | 59 | 61 | 59 | 50 |
| **Age ± SD (year)** |  |  |  |  |  |
| **Mean ± SD** | 53.24±12.55 | 50.05±9.71 | 55.72±13.68 | 50.66±12.95 | 57.02±12.28 |
| **Sex** |  |  |  |  |  |
| **Male (%)** | 102 (44.5%) | 23 (39.0%) | 26 (42.6%) | 30 (50.8%) | 23 (46.0%) |
| **Female (%)** | 127 (55.5%) | 36 (61.0%) | 35 (57.4%) | 29 (49.2%) | 27 (54.0%) |
| **Underlying diseases (%)** |  |  |  |  |  |
| **Allergy** | 14 (6.1%) | 5 (8.5%) | 5 (8.2%) | 3 (5.1%) | 1 (2.0%) |
| **Cardiovascular diseases** | 4 (1.7%) | 3 (5.1%) | 1 (1.6%) | 0 (0.0%) | 0 (0.0%) |
| **Diabetes Mellitus** | 19 (8.3%) | 2 (3.4%) | 8 (13.1%) | 5 (8.5%) | 4 (8.0%) |
| **Dyslipidemia** | 46 (20.1%) | 6 (10.2%) | 17 (27.9%) | 10 (16.9%) | 13 (26.0%) |
| **Hypertension** | 46 (20.1%) | 9 (15.3%) | 17 (27.9%) | 7 (11.9%) | 13 (26.0%) |
| **Thyroid** | 4 (1.7%) | 1 (1.7%) | 3 (4.9%) | 0 (0.0%) | 0 (0.0%) |
| **Others** | 20 (8.7%) | 7 (11.9%) | 10 (16.4%) | 3 (5.1%) | 0 (0.0%) |
| **Follow-up Time** |  |  |  |  |  |
| **Second visit** |  |  |  |  |  |
| **Mean (range) day** | 14.7<br>(12-20) | 14.6<br>(14-20) | 16.0<br>(14-19) | 14.1<br>(13-15) | 14.0<br>(12-21) |
| **Third visit** |  |  |  |  |  |
| **Mean (range) day** | 29.9<br>(21-39) | 28.9<br>(28-36) | 29.9<br>(26-39) | 28.2<br>(26-30) | 33.2<br>(21-35) |
| **Fourth visit** |  |  |  |  |  |
| **Mean (range) day** | 94.5<br>(89-114) | 90.3<br>(90-92) | 102.8<br>(93-114) | 91.8<br>(89-100) | 94.4<br>(90-104) |

### Immunogenicity assessment

#### Total RBD Ig and anti-RBD IgG

There were no differences in total RBD Ig and anti-RBD IgG between four groups at baseline, with geometric mean titer (GMT) 244 U/mL and 50.2 BAU/mL, respectively. Most participants had a very low antibody titer at 5–7 months after completing the second AZD1222 vaccination. However, 14 days after boost, total RBD Ig levels were significantly elevated to 2069, 13651, 28172, and 25824 U/mL in the AZD1222, BNT162b2, mRNA-1273, and half-dose groups, respectively (Figure 2A). Comparable trends with anti-RBD IgG levels were observed with GMT 268, 2285, 3864, and 3976 BAU/mL (Figure 2B). It should be noted that the concentration of total RBD Ig decreased to 1303, 3974, 5516, and 6120 U/mL 90 days after the third dose in each group, respectively. The mRNA-1273 vaccine produced the highest immune response among all booster groups, followed by BNT162b2 and AZD1222, respectively. In comparison, the GMT of total RBD Ig and IgG against RBD between full-and half-dose mRNA-1273 were comparable at both 14 and 90 days (Supplementary Table 1). Anti-N IgG levels were also measured. In this study, the anti-N IgG results showed that all participants were seronegative (cut off 1.4) to anti-N IgG at each visit (Supplementary Figure 1).

**Figure 2:**
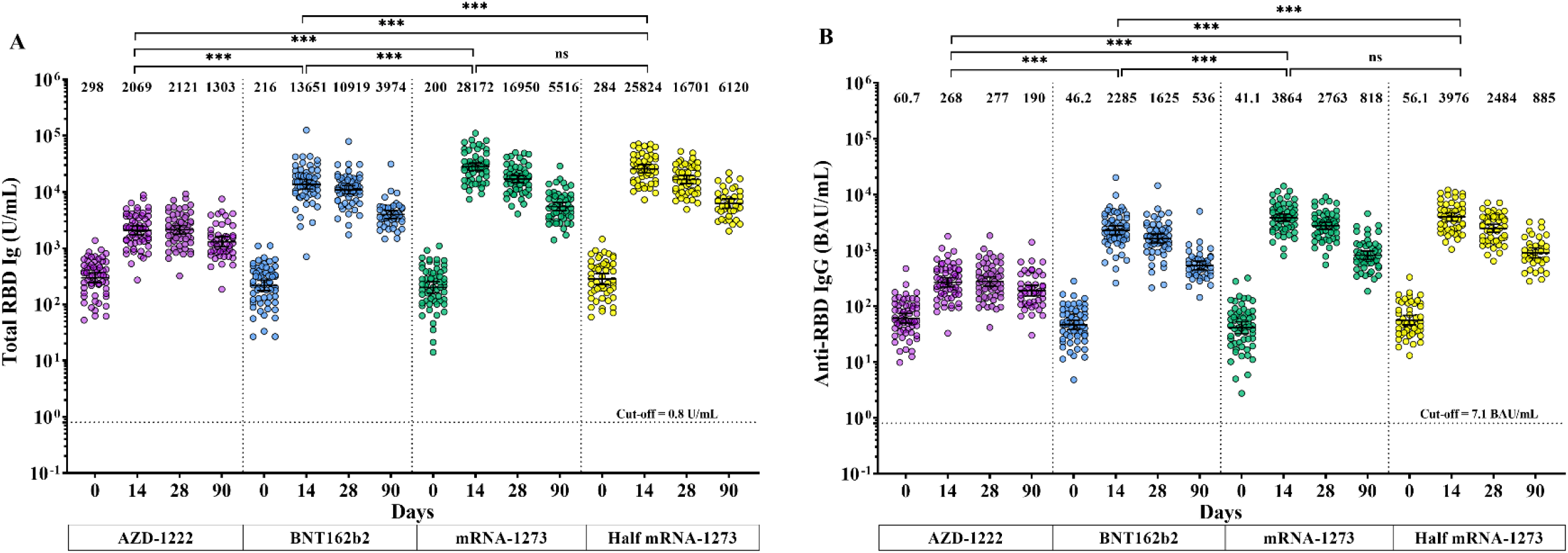
Measurement of the SARS-CoV-2-specific binding antibody. (A) shows the total RBD Ig (U/mL). (B) displays the anti-RBD IgG (BAU/mL). Each data point represents the level of SARS-CoV-2-specific binding antibody in sera from an individual who completed two doses of AZD1222, followed by the booster vaccine, including AZD1222 (purple), BNT162b2 (blue), mRNA-1273 (green), or half-dose mRNA-1273 (yellow). The error bars indicate GMT and 95% confidence intervals (95% CI). The dotted lines designate the cut-off values. ns indicates that there is no statistical difference; **, p<0.01, ***, *p*<0.001.

#### Surrogate virus neutralization specific delta and Omicron variants

The sVNT against the Delta and Omicron variants was performed randomly in half of the participants of each group. A subset of the sample was tested for Surrogate virus neutralization (sVNT) in the pre-booster. At 28 days after boosters, the median percentage inhibition to the Delta variant was significantly elevated to 93.5% in AZD1222, 96.9% in BNT162b2, 97.0% in mRNA-1273 and 97.9% in half dose mRNA-1273 (Figure 3A). While a lower percentage of inhibition was observed against Omicron variants with 15.0%, 67.1%, 64.6%, and 65.7% in groups AZD1222, BNT162b2, mRNA-1273, and half-dose mRNA-1273, respectively (Figure 3B). These results suggested that heterologous boosted individuals achieved a higher level of NAb against the Omicron variant than homologous boosted individuals. On day 90 after vaccination, the median percentage of inhibition to the Delta variant decreased slightly to 78.0% in the AZD1222-boosted group, while the other groups remained >97% inhibition. In contrast to the Omicron variant, the median percentage inhibition was reduced to 9.0% for AZD1222, 33.2% for BNT162b2, 35.5% for mRNA-1273, and 38.7% for half-dose groups of mRNA-1273, respectively. Correspondingly, the neutralizing activity against the Omicron variant was significantly lower than that against the Delta variant (Figure 3C). The heterologous AZD1222/mRNA prime-boosted achieved higher detectable neutralization activities against omicron than the homologous boost at both the 28-day and 90-day timepoint (Supplementary Table 2).

**Figure 3:**
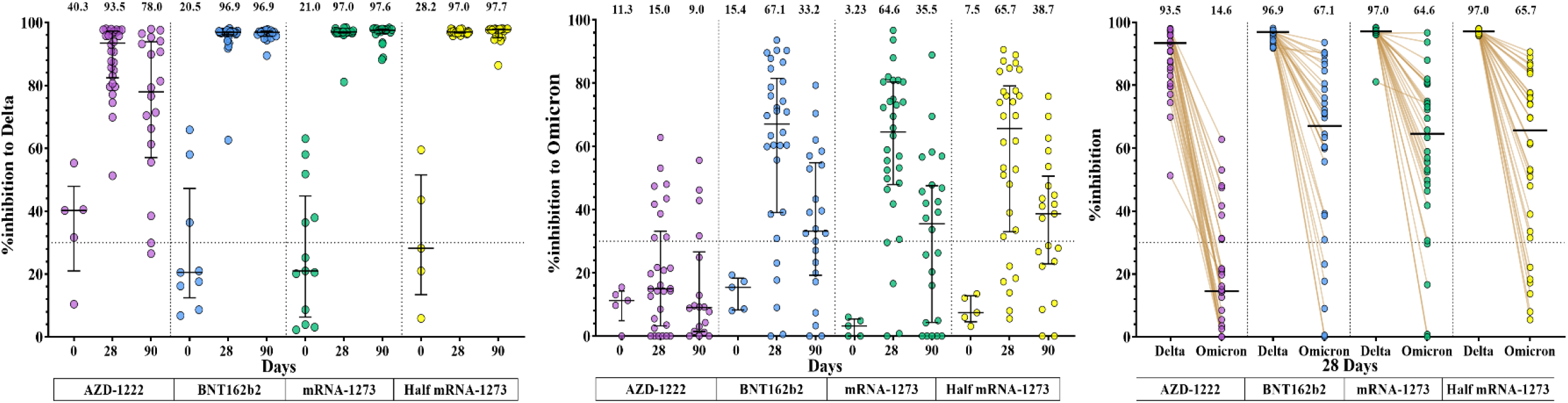
Neutralizing activities of delta and omicron variants compared between pre-and post-boost using a surrogate virus neutralization test. A subset of samples was randomly selected to test for sVNT, including sera collected at baseline and sera collected 28 days after boost (n=30/group). **(A)** Neutralizing activities against the SARS-CoV-2 Delta variant and **(B)** Neutralizing activities against the SARS-CoV-2 Omicron variant (BA.1) were shown as %inhibition. **(C)** Comparison of the neutralizing activity between the Delta and Omicron variants after booster vaccination. Median values with interquartile ranges (IQRs) are denoted as horizontal bars. Horizontal bars indicate the median. Dotted lines designate cutoff values (30%).

#### Focus reduction neutralization test

We determined the cross-neutralizing activity of NAbs against the BA.1 and BA.2 Omicron variants after booster vaccination using the Focus reduction neutralization assay (FRNT50) in a subset of samples. Most participants showed undetectable Nabs on day 0 with 75.0%, 100%, 45%, and 60% in each group. The GMT of NAbs against BA.1 was 32.2 (20.1-51.6), 166 (114-243), 548 (415-723), and 396 (275-571) 28 days after receiving AZD1222, BNT162b2, mRNA-1273, and half-dose mRNA-1273, respectively (Figure 4A). Similarly, GMTs of Nabs against BA.2 were 45.6 (28.8-72.3), 248 (179-342), 324 (214-492), and 224 (156-322), respectively (Figure 4B). Then, 90 days post-vaccination, the GMT levels decreased to 21.6 (13.5-34.8), 87.0 (54.5-139), 141(89.6-222), 119 (78.5-181) for BA.1 and 32.6 (22.0-48.3), 73.8 (56.1-97.2), 139 (85.6-226), 111 (75-163) for BA.2 in group, respectively. The NAb titers to BA.1 were comparable to BA.2 (Figure 4C-4D). Furthermore, higher activity was observed to neutralize the omicron variant in the heterologous mRNA boosted than in homologous boosted individuals.

**Figure 4.**
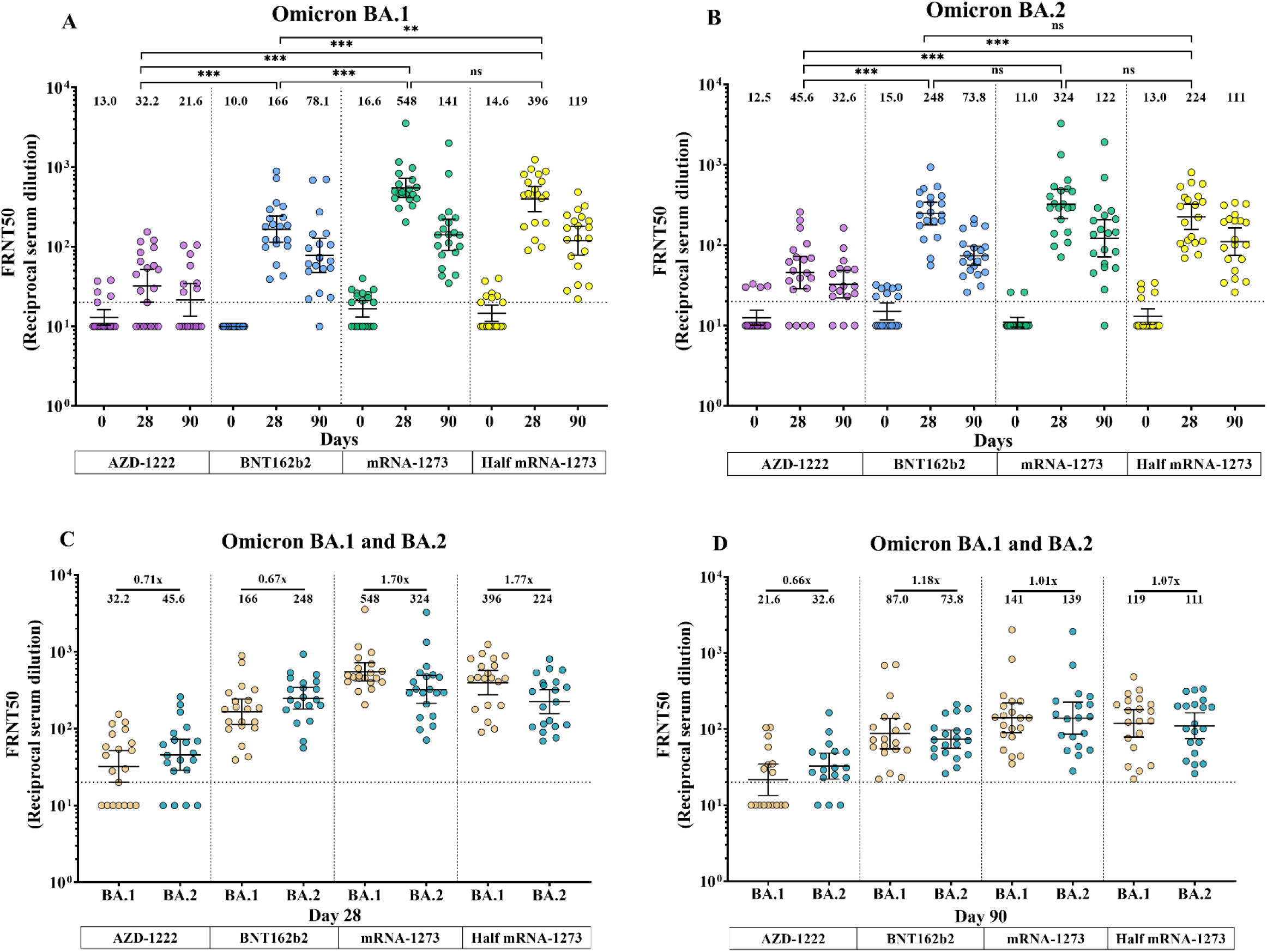
Neutralizing antibody titers against Omicron BA.1 and BA.2 before and after booster vaccination. Neutralization of SARS-CoV-2 Omicron BA.1 and BA.2 was measured using the focus reduction neutralization test (FRNT50). **(A)** Neutralization against the BA.1 omicron variant and **(B)** Neutralization against the BA.2 omicron variant. **(C)** Comparison of NAb titers against BA.1 and BA.2 on 28 days after the booster dose. **(D)** Comparison of NAb titers against BA.1 and BA.2 on 90 days after the booster dose. Each data point represents an individual who received a booster vaccine, including the viral vector vaccine, AZD1222 (purple), the mRNA vaccine, BNT162b2 (blue), mRNA-1273 (green), or half-dose mRNA-1273 (yellow). The error bars present GMT and the 95% confidence interval (CI). Values below the limit were substituted with a titer of 10. ns indicates that there is no statistical difference; **, *p*<0.01, ***, *p*<0.001.

#### Total interferon-gamma response

The IFN-γ release assay was used to measure the presence of T cell responses, for both CD4+ and CD8+ T cells. On day 0, the total IFN-γ level produced by CD4+/CD4+ and CD8+ was less observed in 29/116 (25.0%) and 43/116 (37.1%) of all participants. Conversely, the seropositivity rate for Ag1 CD4+ rapidly increased to 79.3% for BNT162b2, 89.3% for mRNA-1273, 89.3% for the half-dose group on day 14, while the seropositivity rate for Ag2 CD4+ and CD8 + was 86.2%, 89.3% and 92.9%, respectively. Then it slightly decreased at 28 days after boosted (Figure 5A and 5B). The highest seropositivity rate in AZD1222-boosted was 37.9% and 69.0%, indicating that AZD1222-boosted produced a lower T cell response than the mRNA-boosted. Moreover, the persistence of T cell response was observed among mRNA-boosted individuals on day 90 after vaccination, but not in AZD1222-boosted individuals. Consequently, heterologous mRNA boost exhibited a stronger positive impact on T-cell responses than the homologous booster in individuals previously vaccinated with AZD1222. The levels of IFN-γ among all groups are summarized in Supplementary Table 3.

**Figure 5.**
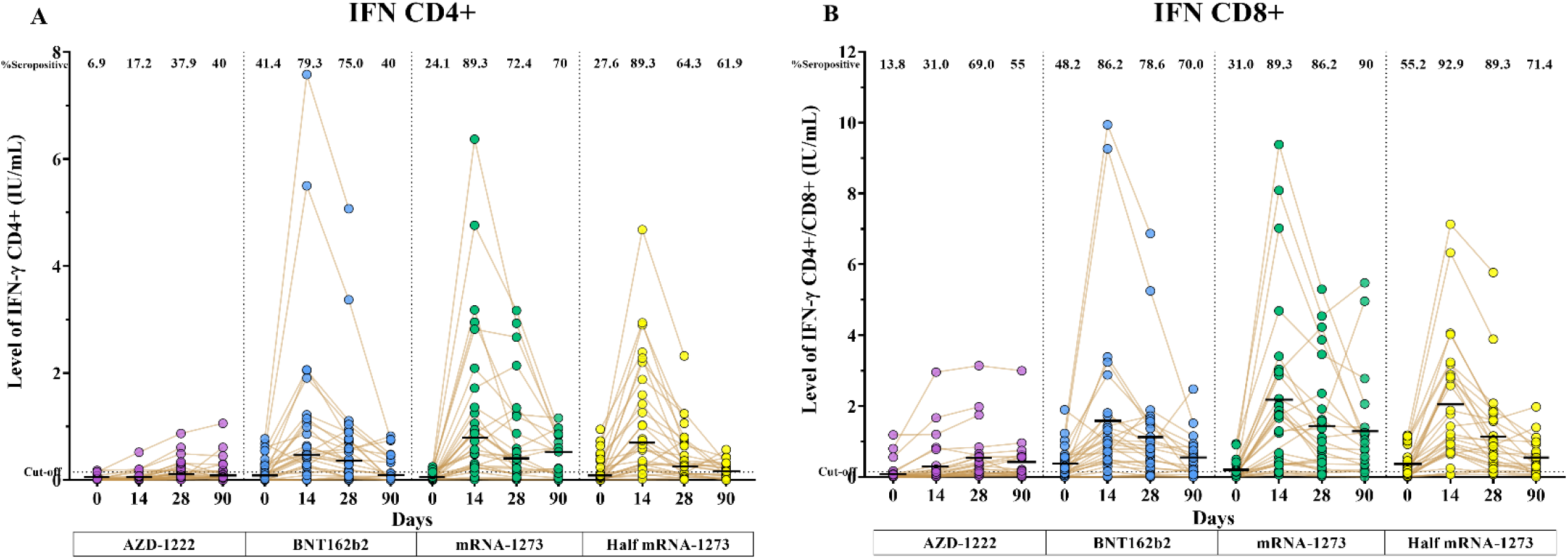
Total CD4+ and CD8+ T cell-producing IFN-γ assay. The QFN IFN-γ ELISA measured levels on days 0, 14, 28, and 90. **(A)** shows the IFN-γ produced by CD4+ (stimulated with CD4+specific epitope RBD from RBD (Ag1)) and (**B**) shows the IFN-γ produced by CD4+ and CD8+ T cell (stimulated with CD4+/CD8+ specific epitope derived from S1 and S2 subunits (Ag2)). Each data point represents an individual who received a booster vaccine, including the viral vector vaccine, AZD1222 (purple), the mRNA vaccine, BNT162b2 (blue), mRNA-1273 (green), or half-dose mRNA-1273 (yellow). Horizontal bars indicate the median. The dotted lines designate the cut-off values.

#### Reactogenicity assessment

Reactogenicities following the booster vaccination were as follows: the most common local vaccine reaction in the first seven days was injection site pain while the most common systemic reaction was myalgia (Figure 6A–6D). Comparison of AEs between groups revealed that the injection site pain, myalgia, and chilling were lower in individuals receiving AZD1222 than in other vaccines (Supplementary Figure 2A-C). However, most incidences of AE were reported as mild-to-moderate in severity and resolved a few days after vaccination. Furthermore, the local reactions AE was less frequent in half-dose mRNA-1273 than in full dose included injection site pain (98.3% vs 86.0%), and swelling (20.8% vs. 30.0%). During the study period, no serious adverse effects were observed among all groups. The summarize data of all AE were shown in Supplementary Table 4.

**Figure 6.**
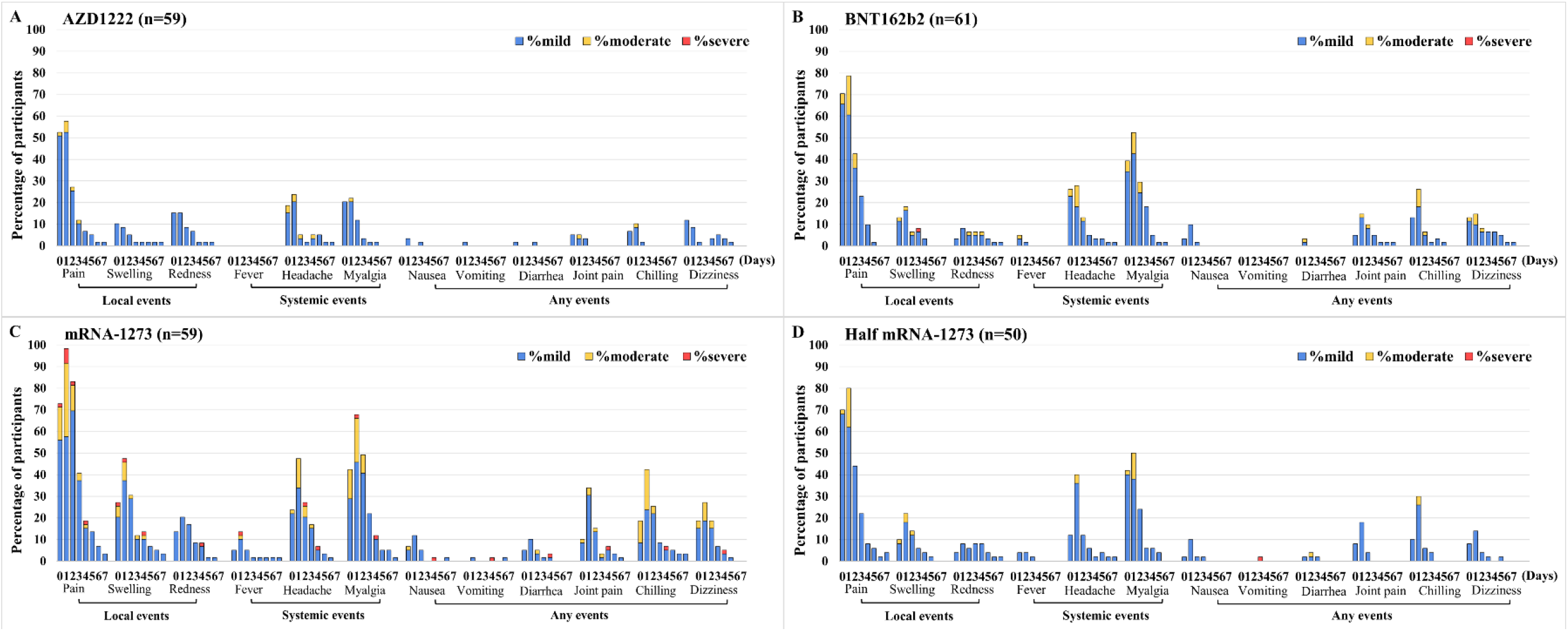
Reactogenicity and severity of local and systemic reactions during the first seven days following vaccination with a booster dose of COVID-19 vaccines. The booster vaccine included the viral vector vaccine – AZD1222 **(A)**, mRNA vaccine – BNT162b2 **(B)**, mRNA vaccine - mRNA-1273 (**C**), and the half dose mRNA-1273 (**D)**. The percentage of participants who reported adverse events is presented on the Y axis. Swelling and redness were classified by measuring the diameter area as mild (<5 cm), moderate (5 to <10 cm), and severe (≥10 cm). Fever was graded as mild (38°C to <38.5°C), moderate (38.5°C to <39°C), and severe (≥39°C). The other events were classified as mild (no impact on regular activity)/ moderate (some limitation of daily activity), and severe (unable to perform or prevented daily activity).

## Discussion

Herein, we report quantifications of total RBD Ig, anti-RBD IgG, Nab, and total IFN-γ levels after homologous and heterologous booster in participants who received two doses of AZD1222 vaccine. We also show the reactogenicity profile of all vaccine boosters. We determined there was a significant increase in total Ig, anti-RBD IgG, neutralizing antibody, and cellular responses 14-28 days after the booster shot. The decay of the immune response was observed 90 days after the initial boosted for all study arms. At 7 days after vaccination, all COVID-19 vaccines administered as a booster dose had acceptable reactogenicity with mild to moderate AE and generally resolved within a few days after vaccination. AE data indicated that homologous and heterologous vaccine boosters presented a good safety profile in all healthy adults, consistent with the previous study of third homologous and heterologous boosters in individuals primed with the AZD-1222 or BNT162b2 vaccines (Flaxman et al., 2021, Munro et al., 2021).

Our findings revealed that following the first primary series of AZD1222, a waning immunity was evidenced 5-7 months after vaccination. After the booster was implemented with both homologous and heterologous vaccines, antibody binding levels increased significantly compared to baseline. Half-dose mRNA-1273 elicited total Ig and IgG RBD responses equal to the full-dose group, which is consistent with the previous observation of mRNA-1273 booster in participants primed with two-doses of mRNA-1273 (Choi et al., 2021). Furthermore, individuals boosted with full and half doses of mRNA-1273 possessed the highest levels of total RBD Ig and anti-RBD IgG compared to those boosted with BNT162b2 and AZD1222. According to the COV-BOOST study, the magnitude of anti-spike IgG in individuals receiving a heterologous booster was significantly higher than the homologous booster (Munro et al., 2021). In addition, similar results have been reported previously regarding the heterologous third dose in individuals receiving triple inactivated or triple mRNA vaccine (Gagne et al., 2022, Liu et al., 2022, Wang et al., 2022). Our results may suggest that a single shot heterologous booster could further enhance the immune response by stimulating the memory B-cell response (Goel et al., 2021).

In accordance with those recent report from the UK, we found that NAbs after 5-7 months of AZD1222-primed individuals rapidly waned to near or below the detection limit and were barely neutralized against Delta and Omicron variants (Wu et al., 2022). Herein, we observed that the heterologous mRNA-based booster, but not the AZD1222-boosted, effectively produced a high level of cross-neutralization against SARS-CoV-2 variants 28 days after the booster dose. The breadth and cross-reactivity of NAbs after the third dose may be related to increasing memory B cell affinity maturation through extensive somatic hypermutation like those observed in a previous mRNA-boosted study (Paschold et al., 2022). However, reductions in neutralizing activity against Omicron variants were found compared with those against Delta variants, in both BNT162b2-primed and CoronaVac-primed indivudals (Assawakosri et al., 2022, Pérez-Then et al., 2022, Planas et al., 2022). Although the BA.2 sublineage was documented to have different viral characteristics than BA.1. Our findings revealed that the neutralizing activity against the BA.1 and BA.2 variants was comparable. These results are consistent with previous studies indicating that BA.2 was only 1.4-fold lower in NAbs than the BA.1 variant after BNT62b2 booster in BNT162b2-primed and previously infected individuals (Chen et al., 2022, Yu et al., 2022). Our finding indicated that mRNA vaccines encoding the ancestral strain-spike could induce a potent cross-neutralizing antibody against Delta, Omicron, both BA.1 and BA.2 subvariants.

Since the NAbs are the leading correlation with protection against COVID-19 infection (Khoury et al., 2021). In our cohort, individuals boosted with mRNA possessed high levels of NAbs titer against BA.1 and BA.2 subvariants. The high NAb titers observed in this study were related to the increase in VE protection against symptomatic disease caused by the Omicron and Delta variants after mRNA booster vaccination among the AZD1222-primed individuals (Andrews et al., 2022). Although >90% of all participants had detectable neutralizing antibody responses after 90 days of booster vaccination, the reduction in NAbs titers against the Omicron variant was evident. Similarly, VE data substantially decreased from 70.1% to 60.9% in mRNA-1273-boosted and 62.4% to 39.6% in BNT162b2-boosted after 10 or more weeks of booster (Andrews et al., 2022). Since there is an evident benefit of a fourth dose seven months after the third dose (Munro et al., 2022), the result showed that the fourth booster elicited a high level of anti-spike IgG in individuals receiving AZD1222/mRNA prime boosters. The additional fourth dose should be optional for those who received the AZD1222 as the third dose due to the lower antibody response in this group.

Besides humoral immune responses, vaccine-induced T cell responses have a potential role in cross-recognizing Alpha, Beta, Delta, and Omicron variants even if using a different vaccination regimen (Gao et al., 2022, Jarjour et al., 2021, Tarke et al., 2021). Because class I and II spike epitopes were 70-80% conserved, leading to the preservation of CD4+ and CD8+ T cell responses (Tarke et al., 2021). Thus, the CD4 + and CD8+ T cells responses have remained stable primarily in frequency and magnitude against all variants of concern. Our study showed the CD4+ and CD8+ T cell-producing IFN-γ were rapidly restored after 14 days of the heterologous booster vaccination. Additionally, >70% of the mRNA-boosted individuals demonstrated seropositive for T cell-producing IFN-γ response after 90 days post-vaccination but showed only 50% seropositive for T cell-producing IFN-γ response in AZD1222-boosted group. Beyond humoral responses, the heterologous booster could enhance the adaptive T-cell response, and the T-cell response may play an essential role as a second-level cross-protection against other upcoming variants.

Potential limitations of our study include may be attributed to the loss of some participants during follow-up. Moreover, the phenotype of CD4+ and CD 8+ T cells was not examined after booster vaccination. Additional studies of 6-month and longer duration are needed to address the long-term durability of the immune response to booster vaccination.

In conclusion, the heterologous mRNA vaccine has an acceptable safety profile and could significantly enhance humoral and cellular responses after vaccination in AZD1222-primed individuals. Additionally, the booster with full-dose and half-dose mRNA-1273 exhibited the highest humoral and cellular immune responses at 90 days after the booster dose, followed by BNT162b2 and AZD1222. Receiving a heterologous booster dose could provide immune protection against new emerging SARS-CoV-2 variants.

## Data Availability

All data produced in the present work are contained in the manuscript.

## Ethics approval and consent to participate

The study protocol was approved by the Institutional Review Board of the Faculty of Medicine of Chulalongkorn University (IRB numbers 690/64). The study was registered with the Thai Clinical Trials Registry (TCTR 20210910002).

## Acknowledgments

The authors are grateful for the Center of Excellence in Clinical Virology staff and all the participants for helping and supporting this project. We also thank the Ministry of Public Health, Chulabhorn Royal Academy, and Zullig pharma for providing the vaccines for this study.

