## Supplementary material for "Persistence of immunity against omicron BA.1 and BA.2 following homologous and heterologous COVID-19 booster vaccines in healthy adults after a two-doses AZD1222 vaccination": Supplementary_Data.docx

**Supplementary Table 1.** Value and statistical analysis of GMT (with 95% confidence intervals) of total RBD Ig and anti-RBD IgG.

|  | **AZD1222**  **(I)** | **BNT162b2**  **(II)** | **mRNA-1273**  **(III)** | **Half mRNA-1273**  **(IV)** | **Kruskal-Wallis** | **Pair-wise analysis (Mann Whitney)** | | | | | |
| --- | --- | --- | --- | --- | --- | --- | --- | --- | --- | --- | --- |
|  |  |  |  |  |  | **(I) vs (II)** | **(I) vs (III)** | **(I) vs (IV)** | **(II) vs (III)** | **(II) vs (IV)** | **(III) vs (IV)** |
| **SARS-CoV-2 total RBD Ig, (U/ml), GMT 95% CI]** | | | | | | | | | | | |
| Day 0 | 298.4  (254.2-363.1) | 216.1 (174.2-268.1) | 200.3 (159.3-252.0) | 283.7  (227.0-354.7) | 0.020 ^a^ | 0.421 ^b^ | 0.055 | 1.000 ^b^ | 1.000 ^b^ | 0.414 | 0.071 |
| Day 14 | 2069 (1724-2484) | 13651 (11179-16668) | 28172 (24077-32963) | 25824  (21786-30611) | < 0.001 ^a^ | < 0.001 ^b^ | < 0.001 | < 0.001 ^b^ | < 0.001 ^b^ | < 0.001 | 1.000 ^b^ |
| Day 28 | 2121  (1774-2536) | 10919 (9226-12922) | 16950 (14677-19574) | 16701 (14130-19740) | < 0.001 ^a^ | < 0.001 ^b^ | < 0.001 | < 0.001 ^b^ | 0.001 ^b^ | 0.003 | 1.000 ^b^ |
| Day 90 | 1303  (1055-1609) | 3974  (3369-4688) | 5516  (4656-6534) | 6120  (4916-7620) | < 0.001 ^a^ | < 0.001 ^b^ | < 0.001 | < 0.001 ^b^ | 0.072 ^b^ | 0.007 | 1.000 ^b^ |
| **SARS-CoV-2 anti-RBD IgG, (BAU/ml), GMT [95% CI]** | | | | | | | | | | | |
| Day 0 | 60.7 (49.7-74.1) | 46.2 (38.1-56.0) | 41.1 (32.2-52.3) | 56.1  (46.1-68.3) | 0.048 ^a^ | 0.366 ^b^ | 0.048 | 1.000 ^b^ | 1.000 ^b^ | 1.000 | 0.249 ^b^ |
| Day 14 | 268  (221-325) | 2285  (1899-2749) | 3864  (3309-4513) | 3976  (3351-4718) | < 0.001 ^a^ | < 0.001 ^b^ | < 0.001 | < 0.001 ^b^ | < 0.001 ^b^ | < 0.001 | 1.000 ^b^ |
| Day 28 | 277 (230-335) | 1625 (1347-1960) | 2763 (2386-3201) | 2484  (2102-2935) | < 0.001 ^a^ | < 0.001 ^b^ | < 0.001 | < 0.001 ^b^ | < 0.001 ^b^ | 0.005 | 1.000 ^b^ |
| Day 90 | 190  (153-237) | 536  (448-639) | 818  (688-973) | 885  (702-1115) | < 0.001 ^a^ | < 0.001 ^b^ | 0.001 | 0.008 ^b^ | 0.016 ^b^ | 0.002 | 1.000 ^b^ |

All tests were adjusted by Bonferroni correction for multiple comparison tests.

^b^ adjusted for age

**Supplementary Table 2.** The percentage inhibition against wild-type and SARS-CoV-2 variants.

|  | AZD1222 (I) | BNT162b2 (II) | mRNA-1273 (III) | Half-dose mRNA-1273 (IV) |
| --- | --- | --- | --- | --- |
| sVNT— Delta. (GenScript), median (IQR) | | | | |
| Day 0 | 40.3 (21.0-47.9) | 20.5 (12.4-47.2) | 21.0 (6.3-44.9) | 28.2 (13.5-51.6) |
| Day 28 | 93.5 (82.4-97.3) | 96.9 (96.2-97.0) | 97.0 (96.7-97.1) | 97.0 (96.7-97.1) |
| Day 90 | 78.0 (57.0-93.9) | 96.9 (95.6-97.3) | 97.6 (96.5-98.3) | 97.7 (95.2-97.9) |
| sVNT—Omicron. (GenScript), median (IQR) | | | | |
| Day 0 | 11.3 (4.86-14.2) | 15.4 (8.28-18.3) | 3.23 (0.0-5.42) | 7.44 (4.52-12.7) |
| Day 28 | 15.0 (3.25-33.2) | 67.1 (39.1-81.5) | 64.6 (47.9-80.4) | 65.7 (33.0-79.1) |
| Day 90 | 9.0 (1.43-26.6) | 33.2 (19.2-54.8) | 35.5 (4.38-47.6) | 38.7 (22.9-50.6) |

**Supplementary Table 3.** The IFN-γ CD4+/IFN-γ CD4+ and CD8+ levels

|  | AZD1222 (I) | BNT162b2 (II) | mRNA-1273 (III) | Half-dose mRNA-1273 (IV) |
| --- | --- | --- | --- | --- |
| IFN-γ CD4+ T-cell (IU/ml), median (IQR) | | | | |
| Day 0 | 0.010 (0.000-0.025) | 0.100 (0.010-0.300) | 0.040 (0.010-0.145) | 0.070 (0.010-0.300) |
| Day 14 | 0.020 (0.010-0.060) | 0.490 (0.245-1.120) | 0.800 (0.245-1.630) | 0.700 (0.250-1.810) |
| Day 28 | 0.120 (0.050-0.230) | 0.350 (0.110-0.760) | 0.400 (0.125-1.210) | 0.270 (0.124-0.673) |
| Day 90 | 0.095 (0.050-0.263) | 0.455 (0.110-0.820) | 0.530 (0.100-0.855) | 0.175 (0.083-0.308) |
| IFN-γ CD4+/CD8+ T-cell (IU/ml), median (IQR) | | | | |
| Day 0 | 0.020 (0.010-0.055) | 0.175 (0.080-0.545) | 0.060 (0.030-0.185) | 0.215 (0.028-0.485) |
| Day 14 | 0.040 (0.010-0.215) | 0.920 (0.428-1.510) | 1.700 (0.350-2.950) | 1.320 (0.733-2.890) |
| Day 28 | 0.240 (0.115-0.580) | 0.770 (0.235-1.440) | 0.810 (0.315-1.770) | 0.820 (0.363-1.600) |
| Day 90 | 0.165 (0.073-0.565) | 0.310 (0.070-0.810) | 0.875 (0.335-1.360) | 0.340 (0.140-0.878) |

**Supplementary Table 4.** Statistical analysis of reactogenicity data between booster vaccines.

|  | Total | AZD1222 | BNT162b2 | mRNA-1273 | Half mRNA-1273 | Pearson’s χ^2^ | Pair-wise analysis (Fisher’s Exact test) | | | | | |
| --- | --- | --- | --- | --- | --- | --- | --- | --- | --- | --- | --- | --- |
|  |  |  |  |  |  |  | 1 vs 2 | 1 vs 3 | 1 vs 4 | 2 vs 3 | 2 vs 4 | 3 vs 4 |
| N (%) | 229 (100) | 59 (100) | 61 (100) | 59 (100) | 50 (100) |  |  |  |  |  |  |  |
| Injection site pain | 196 (85.6) | 42 (71.2) | 53 (86.9) | 58 (98.3) | 43 (86.0) | < 0.001 | 0.043 | < 0.001 | 0.069 | 0.032 | 1.000 | 0.023 |
| Swelling | 68 (29.7) | 9 (15.3) | 14 (23.0) | 30 (50.8) | 15 (30.0) | < 0.001 | 0.356 | < 0.001 | 0.103 | 0.002 | 0.515 | 0.033 |
| Redness | 36 (15.7) | 10 (16.9) | 6 (9.8) | 14 (23.7) | 6 (12.0) | 0.169 | 0.292 | 0.493 | 0.590 | 0.051 | 0.766 | 0.140 |
| Fever | 13 (5.7) | 0 (0.0) | 3 (4.9) | 8 (13.6) | 2 (4.0) | 0.006 ^a^ | 0.244 | 0.006 | 0.208 | 0.123 | 1.000 | 0.105 |
| Headache | 100 (43.7) | 23 (39.0) | 24 (39.3) | 32 (54.2) | 21 (42.0) | 0.293 | 1.000 | 0.140 | 0.845 | 0.143 | 0.847 | 0.250 |
| Myalgia | 133 (58.1) | 20 (33.9) | 38 (62.3) | 44 (74.6) | 31 (62.0) | < 0.001 | 0.002 | < 0.001 | 0.004 | 0.172 | 1.000 | 0.213 |
| Nausea | 25 (10.9) | 3 (5.1) | 7 (11.5) | 9 (15.3) | 6 (12.0) | 0.350 | 0.323 | 0.125 | 0.296 | 0.599 | 1.000 | 0.782 |
| Vomiting | 5 (2.2) | 1 (1.7) | 0 (0.0) | 3 (5.1) | 1 (2.0) | 0.215 ^a^ | 0.492 | 0.619 | 1.000 | 0.116 | 0.450 | 0.623 |
| Diarrhea | 17 (7.4) | 2 (3.4) | 2 (3.3) | 10 (16.9) | 3 (6.0) | 0.013 | 1.000 | 0.029 | 0.659 | 0.015 | 0.656 | 0.136 |
| Joint pain | 47 (20.5) | 5 (8.5) | 9 (14.8) | 22 (37.3) | 11 (22.0) | 0.001 | 0.396 | < 0.001 | 0.059 | 0.006 | 0.335 | 0.097 |
| Chilling | 73 (31.9) | 10 (16.9) | 20 (32.8) | 27 (45.8) | 16 (32.0) | 0.01 | 0.058 | 0.001 | 0.075 | 0.191 | 1.000 | 0.171 |
| Dizziness | 57 (24.9) | 13 (22.0) | 13 (21.3) | 21 (35.6) | 10 (20.0) | 0.177 | 1.000 | 0.154 | 0.819 | 0.106 | 1.000 | 0.090 |

^a^ Likelihood Ratio is applied due to expected cell count less than 5 more than 20% of cells

**Supplementary Figure 1.** **Measurement of the anti-nucleocapsid IgG.** Level of IgG anti-nucleocapsid (N) of SARS-CoV-2 index (S/C) were measured on days 0, 14, 28, and 90. Serum samples were obtained from participants who completed two doses of the AZD1222, followed by the AZD1222, mRNA vaccine–BNT162b2, mRNA-1273, or half-dose mRNA-1273. Lines represent the median (interquartile range). The dotted lines designate the cut-off values.

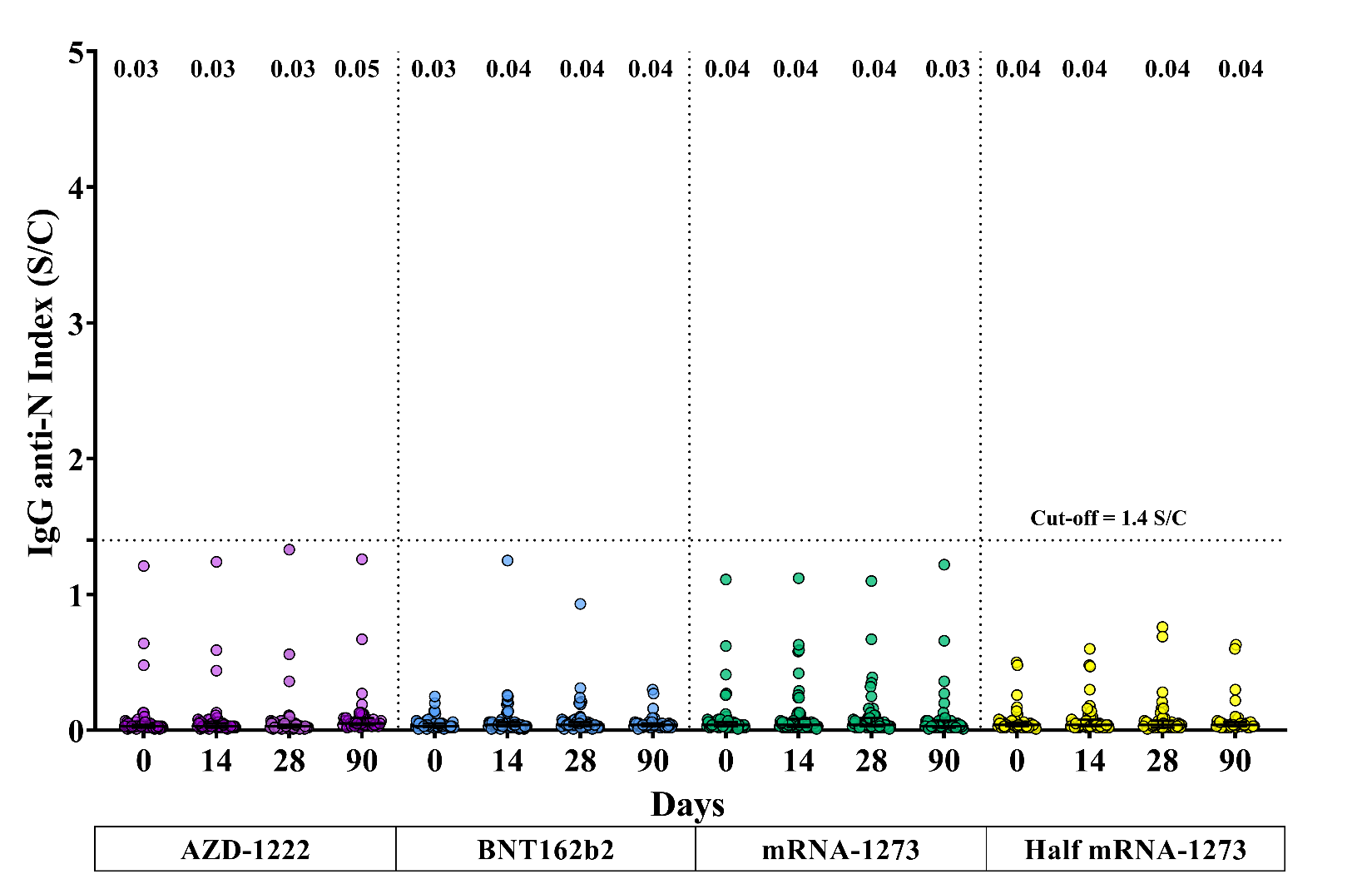

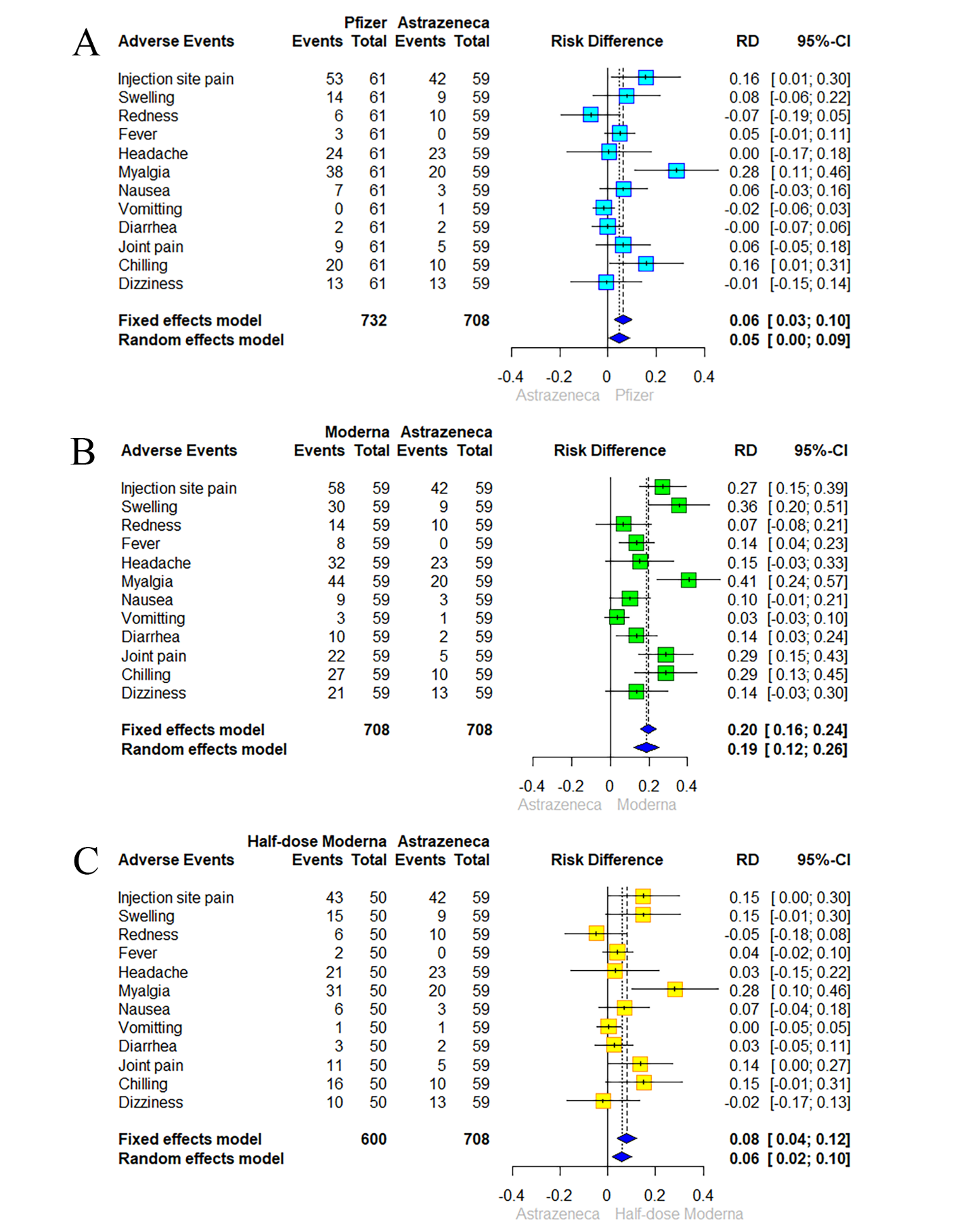
**Supplementary Figure 2A-C. The Forest plot compare the local and systemic adverse events between vaccine groups.** Forest plot presents any grade the solicited local and systemic adverse events (AEs) across 7 days post-boost vaccination and the absolute risk differences comparison between homologous AZD1222 to heterologous BNT162b2, full- and half-dose mRNA1273 in the proportion of participants with 95% confidence intervals.
